# Prevalence and genotype distribution of human papillomavirus (HPV) among adolescent girls and young women in a high HIV burden rural area of South Africa: a cross-sectional survey

**DOI:** 10.64898/2026.09.09.26362609

**Authors:** Maphe Mthembu, Kathy Baisley, Jacob Busang, Nqobile Ngoma, Nonsikelelo Ndlela, Nonhlanhla Okesola, Carina Herbst, Thandeka Khoza, Jaco Dreyer, Lenine Liebenberg, Zaza Ndhlovu, Maryam Shahmanesh

## Abstract

**Background:** Human papillomavirus (HPV) is the primary cause of cervical cancer, with women living with HIV at increased risk of HPV infection and HPV-related disease. Data on HPV prevalence and genotype distribution among adolescent girls and young women (AGYW) in rural South Africa settings remain limited. We estimated HPV prevalence, genotype distribution, and factor associated with HPV infection among AGYW in a high HIV prevalence district of KwaZulu-Natal, South Africa.

**Methods:** This cross-sectional study was nested within the baseline survey of the Thetha Nami population-based trial among young people aged 15-30 years in rural uMkhanyakude District, KwaZulu-Natal. Survey participants were asked to provide dried blood spots for HIV testing and genital swabs for STI testing. For this study, we selected AGYW living with HIV with available vaginal swabs and a frequency-matched sample of AGYW without HIV. HPV genotyping was done using the HPV Direct Flow Chip Kit (Master Diagnostica, Spain). Genotype-specific HPV prevalence was described overall and by HIV status. Logistic regression was used to identify factors associated with HPV infection.

**Results:** Among 187 AGYW with valid HPV results, 90 (48.1%) were living with HIV. Overall HPV prevalence was 69.1% (95% confidence interval (CI)=54.5-80.7%) and high-risk (hr)HPV prevalence was 57.3% (95%CI=44.1-69.6%). HPV prevalence was higher among AGYW living with HIV than among AGYW without HIV (84.6% vs 64.6%, p=0.02). HPV genotype distribution varied by HIV status: HPV35, HPV45 and HPV58 were the most prevalent hrHPV genotypes among AGYW living with HIV, vs HPV52, HPV56 and HPV31 among AGYW without HIV. No HPV16 or HPV18 infections were detected among participants who were age-eligible for the national HPV vaccination programme. Reported condomless sex (adjusted odds ratio (aOR)=2.23, 95%CI=1.06–4.70) and the presence of another sexually transmitted infection (aOR=4.30, 95%CI=1.52–12.20) were associated with HPV infection.

**Conclusions:** AGYW in rural uMkhanyakude experience a high burden of hrHPV infection, especially those living with HIV. Regardless of HPV vaccination status not being confirmed, no HPV 16/18 was detected among vaccine-eligible AGYW. The higher prevalence of non-HPV16/18 hrHPV genotypes covered by the nonavalent vaccine highlights the potential value of vaccines with broader genotype coverage. These findings support expanding HPV screening and cervical cancer prevention programmes in high HIV prevalence settings.

## Introduction

Human papillomavirus (HPV) infection is the primary cause of cervical cancer and a major public health problem in sub-Saharan Africa. Cervical cancer is the 2^nd^ leading cause of cancer-related deaths among women in sub-Saharan Africa (1, 2). HPV is one of the most common sexually transmitted infections (STIs), and is also responsible for anogenital and oropharyngeal cancers in both women and men (3). Most HPV infections are transient and cleared naturally; however, persistent infection with oncogenic, or high-risk, HPV types (hrHPV) significantly increases the risk of cervical intraepithelial neoplasia (CIN) and progression to invasive cervical cancer. South Africa is among the countries greatly burdened by cervical cancer incidence and mortality, with notable regional disparities. Rural areas, such as parts of KwaZulu-Natal, face an even greater burden (2, 4). Contributing factors include socioeconomic challenges, which translate to limited healthcare access, and a high rate of HIV co-infection (5–9).

Studies have shown that women living with HIV (WLWH) are at increased risk of persistent hrHPV infection due to immunosuppression, which impairs viral clearance and increases susceptibility to persistent infections and progression to cervical cancer, leading to an increased incidence of cervical dysplasia and malignancies (10–13). The interplay between HPV and HIV presents a significant public health challenge, particularly in regions with high HIV prevalence such as KwaZulu-Natal (14). Population studies in South Africa have reported HPV prevalence rates as high as 80% among WLWH, with multiple genotype infections being common (7, 15, 16). Despite the well-established association between HPV and HIV, HPV surveillance and genotype characterization remain limited in rural South African populations.

Current HPV vaccines are prophylactic and are most effective in individuals who have not yet been infected with HPV. Therefore, the primary target age for vaccination is before sexual debut. In South Africa, the government rolled out a national school-based vaccination program in 2014 targeting girls aged 9–12 years using the bivalent HPV vaccine Cervarix®, which protects against HPV16 and 18, the cause of 70-80% of cervical cancers (17). However, vaccine uptake in rural communities remains suboptimal due to logistical barriers, literacy, access, vaccine hesitancy, and limited HPV awareness (18, 19).

Cervical cancer screening coverage in South Africa has historically remained suboptimal. However, recent 2024/25 reports suggest modest improvement (38% nationally) (20). Significant disparities persist, with rural women continuing to face geographic, financial, and cultural barriers to accessing screening services (21, 22). Our study uses a unique opportunity provided by a population-representative survey to describe the HPV prevalence and genotype distribution among adolescent girls and young women (AGYW) in the rural uMkhanyakude district, KwaZulu-Natal.

## Methods

### Study population

Our study was nested within the 2022 baseline cross-sectional survey conducted as part of *ThethaNami NgithetheNawe*, cluster-randomised trial (23), and used data collected through this parent study’s survey. For the baseline survey of the trial, the health and demographic surveillance system (HDSS) of the Africa Health Research Institute (AHRI) was used as a sampling frame to select a random sample of 3600 young people aged 15-30 years, stratified by sex and age group (15–19, 20–24, 25–30 years). All AGYW participants were asked to provide STI testing samples, either urine or self-collected vaginal swabs, and a dried blood spot (DBS) for HIV testing. For this nested study, we restricted our sample to AGYW of known HIV status with an available vaginal swab. We selected all AGYW living with HIV who also had available vaginal swab samples and frequency-matched them by age group to an equal number of HIV-negative participants.

### HPV DNA screening and genotyping

After collection, vaginal swabs were stored dry in cryovials at –80 °C. HPV testing for the nested study was conducted 2-3 years after samples were stored. Swabs were thawed and DNA was extracted using the QIAamp DNA Mini Kit (Qiagen, Hilden, Germany) according to the manufacturer’s protocol. Briefly, swab material was eluted in nuclease-free water, lysed with proteinase K, and nucleic acids were bound to silica membrane spin columns. After washing to remove contaminants, DNA was eluted in 50 µL of nuclease-free buffer and quantified using a NanoDrop spectrophotometer (Thermo Fisher Scientific, USA).

For overall HPV screening and genotyping, we used the HPV Direct Flow Chip Kit (Master Diagnostica, Granada, Spain) as per the manufacturer’s instructions. Briefly, the extracted DNA (targeting a total concentration of 100 ng and an A260/A280 purity ratio of 1.8–2.0) was subjected to multiplex PCR targeting the HPV L1 gene, with β-globin included as an internal control of sample adequacy. Amplified products were hybridized to type-specific oligonucleotide probes immobilized on flow-through membrane strips. This kit enables the detection of 35 HPV genotypes (HPV-6, -11, -16, -18, -26, -31, -33, -35, -39, -40, -42, -43, -44/-55, -45, -51, -52, -53, -54, -56, -58, -59, -61, -62/-81, -66, -67, -68, -69, -70, -71, -72, -73, -82 and -84), including the 2 pairs (HPV-44/55 and -62/81) that the assay presents indistinguishable. For comparability with global data, HPV genotypes were classified according to the IARC risk classification, whereby 13 genotypes are defined as high-risk (HPV 16, 18, 31, 33, 35, 39, 45, 51, 52, 56, 58, 59, and 68). Hybridization and washing were automated using the HybriSpot 12 platform®, Vitro Master Diagnostica (Spain) and results were visualized by colorimetric detection. Genotypes were assigned using the automated HybriSoft 2.2.0 R18 (HS12a) software. Positive and negative controls supplied with the kit were included in each run. Samples lacking both HPV signal and β-globin amplification were considered invalid and either repeated or excluded depending on sample availability. Participants were classified as HPV-positive if one or more genotypes were detected.

This combined workflow of DNA extraction using Qiagen technology and HPV genotyping with the Direct Flow Chip Kit was selected for its sensitivity, robustness, and ability to simultaneously detect multiple genotypes, making it suitable for large-scale cohort studies in high-HPV burden settings.

### Statistical methods

Sociodemographic, behavioral, and clinical characteristics were summarized overall and by HIV status, with differences assessed using Fisher’s exact test for categorical variables and the Wilcoxon rank-sum test for continuous variables. HPV prevalence was estimated within age and HIV-status strata using unweighted data. To obtain HPV prevalence estimates representative of the target population, post-stratification weights were calculated based on the age and HIV-status distribution among AGYW in the HDSS. HPV prevalence within each HIV-status group was weighted to reflect the age distribution of the corresponding HIV-status population, and overall HPV prevalence was weighted to reflect the joint age and HIV-status distribution of the population. Overall age-specific prevalence estimates were similarly standardised to the HIV distribution within each age group. Confidence intervals were calculated using Wilson’s method.

Factors associated with any HPV infection were examined using Firth’s penalized logistic regression, because of the small sample size and potential issues with sparse data and separation. All models were adjusted a priori for age group and HIV status. No attempt was made to build a full multivariable model with all potential covariates, because of sample size limitations and to avoid model overfitting. The contribution of individual covariates was assessed using penalized likelihood ratio tests, whereby the full model including the covariates of interest was compared with a constrained model in which the coefficients for those covariates were fixed to zero, in order to retain the covariate in the penalization. To explore the potential impact of South Africa’s national HPV vaccination programme, HPV16/18 prevalence was examined stratified by whether participants were likely to have been offered vaccination, based on their age at the time of the baseline survey in 2022. Participants aged >20 years were classified as likely unvaccinated, while those aged ≤20 years were classified as likely vaccinated..

## Results

Among the 3599 individuals sampled for the *ThethaNami NgithetheNawe* baseline survey, 2721 (75.6%) were found to be still resident and eligible, and 2094 (77.0%) were enrolled. Among 2094 participants, 1131 (54.0%) were AGYW. Of these, 1025 (90.6%) consented to dried blood spot testing and had valid HIV results, of whom 214 (20.9%) were found to be living with HIV. Overall, 779/1025 (76%) of people with valid HIV results consented for curable STI testing (gonorrhea, chlamydia, or trichomoniasis) done in the parent study, and this did not differ much by HIV status. Furthermore, 383/779 (49.9%) AGYW opted for vaginal swab rather than urine collection; this STI sampling was higher among AGYW living with HIV (59.1%, 101/171) compared to those without HIV (46.4%, 282/608). In total, 101 AGYW living with HIV and 101 age-matched AGYW without HIV were selected for HPV testing; 187/202 (92.6%) (90 living with HIV and 97 without HIV) had valid HPV results based on β-globin amplification and were included in this study (Fig. 1, Supplementary Table 1 and 2).

**Figure 1:**
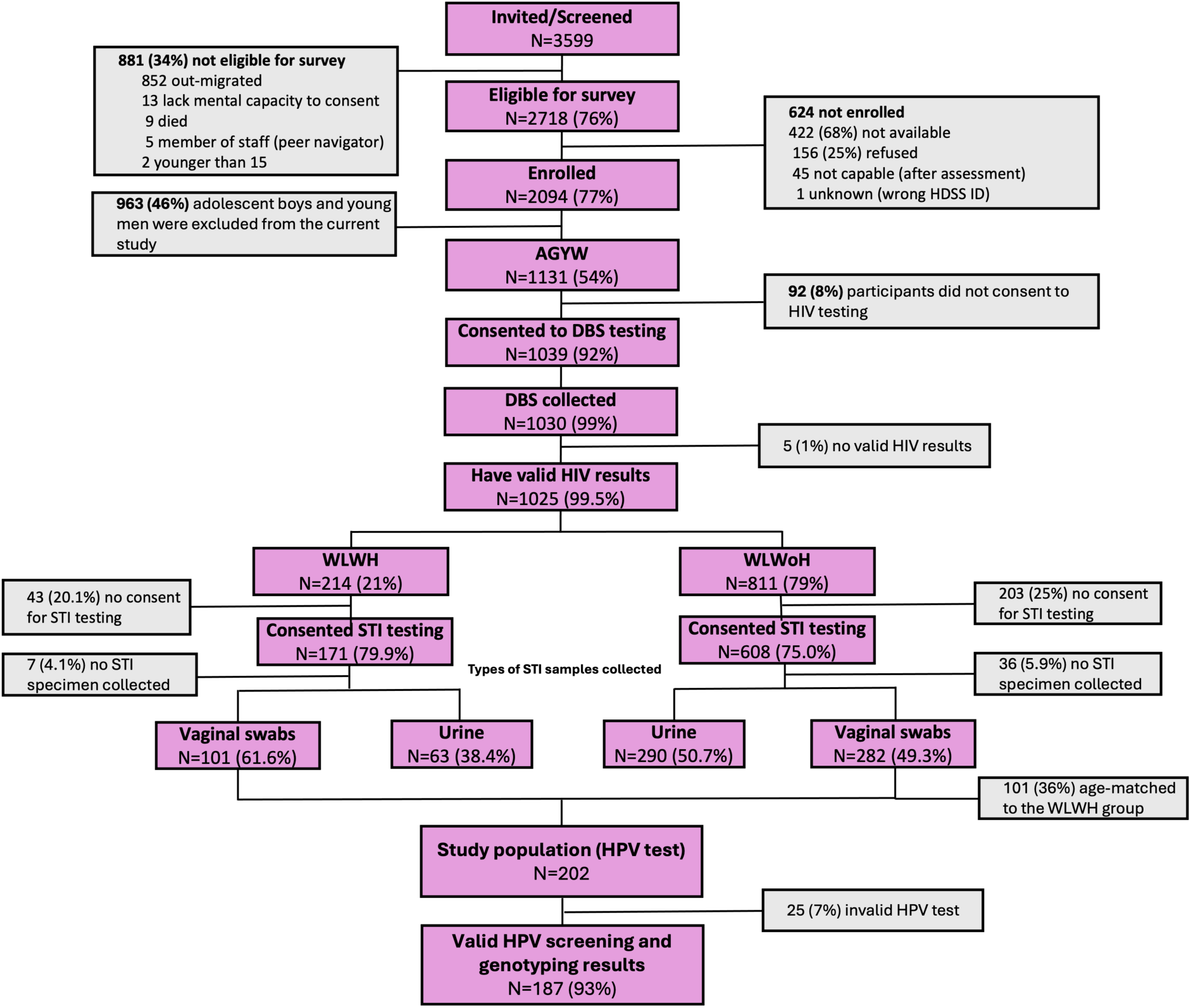
Strategic Selection of Participants from *Thethanami Ngithethenawe* for Inclusion in the Current Study. Flowchart showing participant enrollment at baseline in both the parent study and the current study, and the process leading to the final number of participants screened in the present analysis. N = samples size, WLWH = women living with HIV, WLWoH = women living without

Participant characteristics are shown in Table 1. Median age was 26 years (IQR 22-29). The majority of participants were neither employed nor studying (72.7%). Most participants reported no alcohol use (80.7%) and were non-smokers (96.8%), with no evidence of a difference between AGYW living with HIV and AGYW without HIV (p>0.99 and 0.46, respectively). Food insecurity was reported by nearly half of the participants (47.1%). AGYW living with HIV were more likely to report having at least one sexual partner in the past 12 months (P = 0.05) and the use of contraception (P = 0.01) than AGYW without HIV. There was no evidence of a difference in reported condom use, transactional sex, or the prevalence of co-infection with other STIs between AGYW living with HIV and AGYW without HIV (p = 0.56, 0.19, and >0.99, respectively) (Table 1).

Overall, 154/187 participants tested positive for HPV DNA (weighted prevalence 69.1%, 95%CI=54.5-80.7%). HPV prevalence was higher among AGYW living with HIV than AGYW without HIV (weighted prevalence 84.6% vs 64.6%, respectively, p=0.02). (Table 2). HPV prevalence was also higher among AGYW living with HIV aged under 25 years than their age counterparts without HIV (weighted prevalence 93.1% vs 60.4%, p=0.001). In contrast, among those aged 25-30 years, HPV prevalence did not differ by HIV status (78.2% in both HIV groups, p>0.99). Among AGYW living with HIV, HPV prevalence was higher among those with viral load >200 copies/mL compared with those who were virally suppressed, although the difference was not statistically significant (weighted prevalence 90.2% vs 80.1%, respectively, p=0.20).

HrHPV DNA was detected in 115/187 participants (weighted prevalence 57.3%, 95%CI=44.1-69.6%). The most prevalent hrHPV types were HPV52 (weighted prevalence=14.6%), HPV56 (12.6%) and HPV31 (11.6%) (Table 3, Figure 2). The most prevalent lrHPV types were HPV44/55 (weighted prevalence=14.6%) and HPV62/81 (13.5%). HPV genotype distribution differed by HIV status. Among AGYW living with HIV, the most frequently detected hrHPV types were HPV-35, HPV-45, and HPV-58 (Table 3, Figure 3). Among AGYW without HIV, the most frequently detected hrHPV types were HPV-52, HPV-56, and HPV-31.

**Figure 2.**
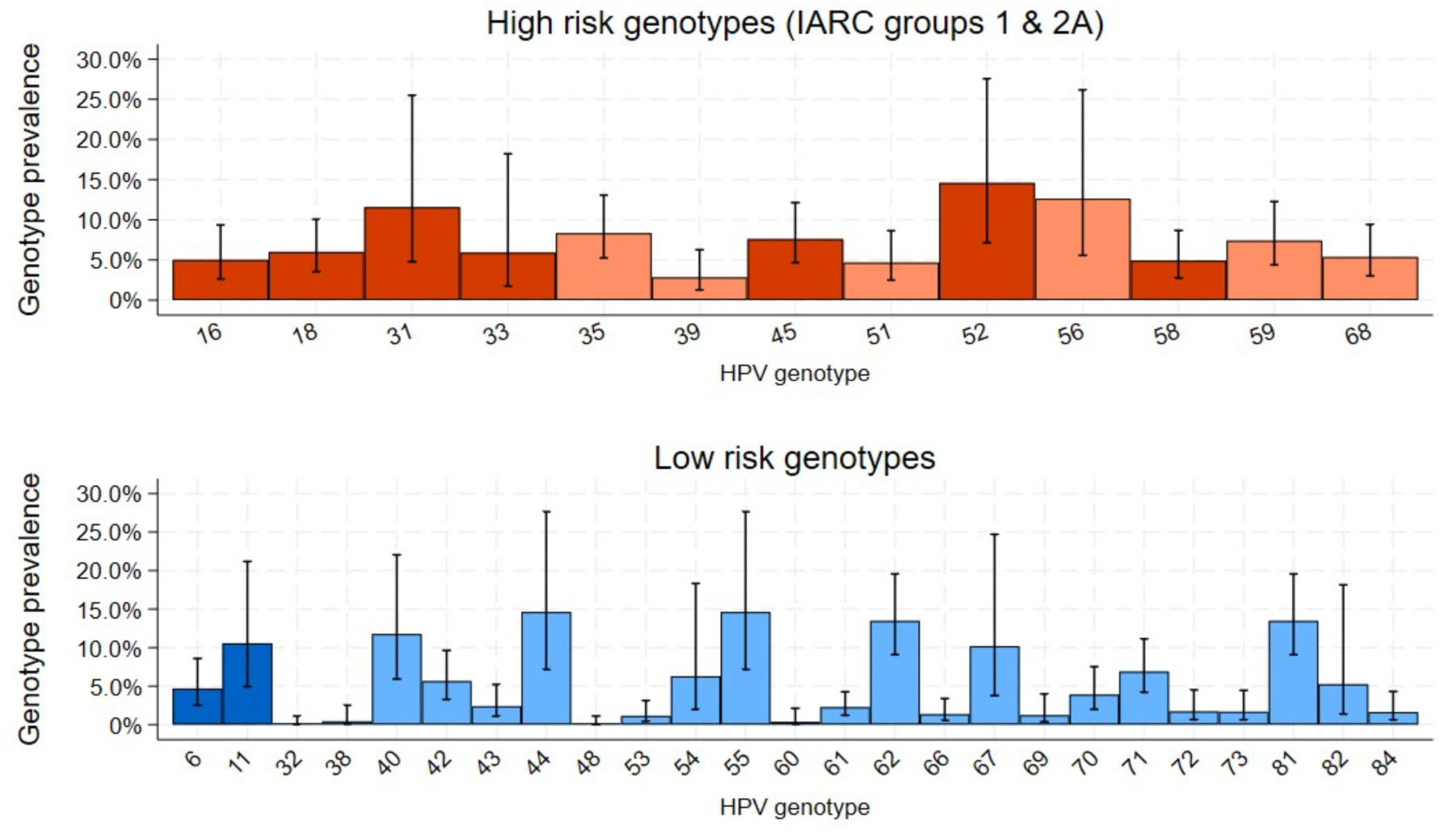
Estimated population prevalence of HPV genotype-specific DNA among adolescent girls and young women aged 15-30 years in rural South Africa. Dark orange/dark blue bars are genotypes covered by 9-valent vaccine. Bars represent 95% confidence intervals.

**Figure 3.**
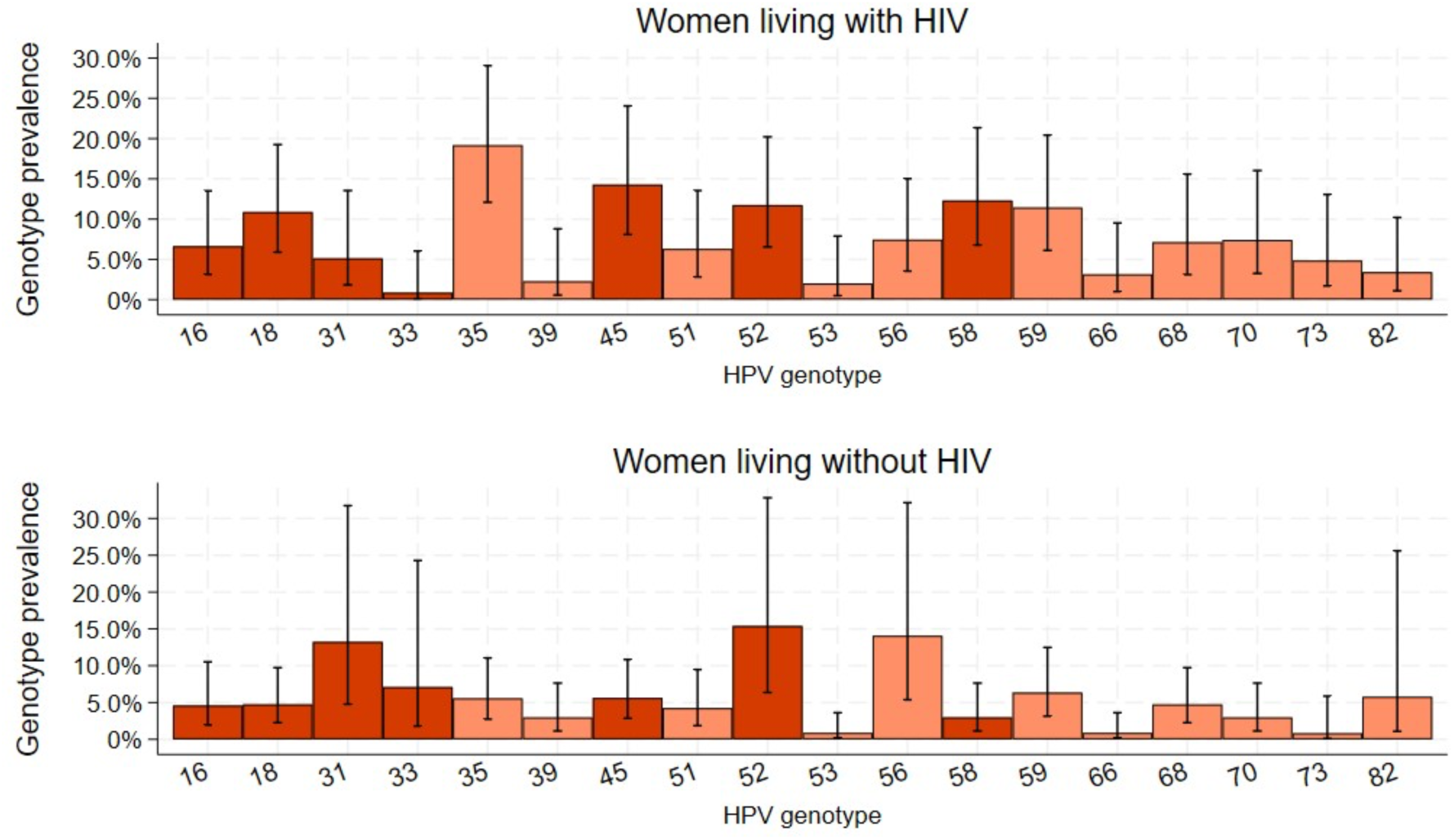
Estimated population prevalence of high-risk HPV genotype-specific DNA among adolescent girls and young women aged 15-30 years in rural South Africa, stratified by HIV status. Dark orange/dark blue bars are genotypes covered by 9-valent vaccine. Bars represent 95% confidence intervals.

The overall weighted prevalence of the 7 hrHPV genotypes targeted by the nonavalent vaccine was 41.0% (95%CI=29.2-54.1%) and was similar between AGYW living with HIV and AGYW without HIV (weighted prevalence 45.3% vs 39.9%, respectively, p=0.60). The overall weighted prevalence of HPV16/18 was 10.0% (95%CI=6.4-15.4%), and was higher among AGYW living with HIV than AGYW without HIV (17.3% vs 7.8%, respectively, p=0.04). However, there were no HPV 16/18 infections among AGYW aged <20 years (Table 2). The prevalence of HPV genotypes for which Cervarix has been reported to provide some cross-protection (HPV-33, 33, and 45) (24) was higher in the likely vaccinated than unvaccinated group (weighted prevalence 21.9% vs 16.3%, respectively), although the difference was not statistically significant (p=0.61). The 7 hrHPV genotypes targeted by the nonavalent vaccine (HPV16, 18, 31, 33, 45, 52, and 58) were common in both likely vaccinated and unvaccinated groups (41.7% vs. 40.6%, respectively).

After adjusting for age group and HIV status, AGYW reporting condomless sex in the past 3 months had over two times the odds of HPV infection as those reporting no condomless sex (adjusted odds ratio (aOR)=2.23; 95%CI=1.06–4.70; p=0.03) (Table 4). HPV infection was also associated with having had more partners in the past 12 months. Similarly, AGYW diagnosed with at least one other curable STIs had more than four times higher odds of HPV infection compared with those without an STI (aOR=4.30; 95%CI=1.52–12.20; p=0.002). AGYW who reported alcohol use in the past month had lower odds of HPV infection than those who did not report alcohol use (aOR=0.40; 95%CI=0.17–0.94; p=0.04).

## Discussion

Data on HPV epidemiology among AGYW in rural South African communities remain limited despite the high burden of both HIV and cervical cancer. In this population-based study from uMkhanyakude District, KwaZulu-Natal, an area with a high HIV burden, we found an HPV DNA prevalence of 69.1% and an hrHPV DNA prevalence of 57.3%. HPV infection was common among both AGYW living with and without HIV, although prevalence was highest among AGYW living with HIV who were not virally suppressed. We also observed differences in genotype distribution by HIV status and found no HPV16/18 infections among participants likely to have been eligible for HPV vaccination.

The overall prevalence of HPV in our study was in the higher range of that reported in many studies from South Africa, which ranged from 33-65% (5, 7, 9, 16, 25). Furthermore, HPV prevalence was significantly lower among HIV-negative participants (65%) than among participants living with HIV (85%). Compared with HIV-negative AGYW, those living with HIV who were virally suppressed (viral load <200 copies/mL) also had a higher HPV prevalence (80%). HPV prevalence was similarly high among virally suppressed (80%) and unsuppressed (90%) AGYW living with HIV, suggesting that viral suppression alone may not fully mitigate the increased risk of HPV infection in this population. This pattern has been observed elsewhere and supports the focus on cervical screening in WHLH (11, 26).

Among AGYW living with HIV, HPV-35 and HPV-45 were the most prevalent hrHPV genotypes, consistent with reports from several studies conducted among women living with HIV in sub-Saharan Africa. A different genotype profile was observed among AGYW without HIV, with HPV-52, HPV-56 and HPV-31 being the most prevalent genotypes. Similar observations have been made elsewhere, where HPV-35 and HPV-45 are disproportionately represented among women living with HIV compared with the general population (3, 25, 27). These observed differences in hrHPV genotype distribution by HIV status highlight the importance of considering HIV status when interpreting HPV epidemiological data in high HIV prevalence settings. Comparisons between studies should take differences in HIV prevalence into account. Although our study was not designed to evaluate vaccination or screening policies, these differences may have implications for modelling the potential impact of HPV vaccination and for planning cervical cancer screening programmes in populations with a high burden of HIV.

No HPV16 or HPV18 infections were detected among AGYW living with HIV or without HIV, in the age group likely to have been eligible for HPV vaccination (<20 years old). Although vaccination records were not available and the sample size was limited, this finding is compatible with the reductions in vaccine-type HPV infections reported following the introduction of national vaccination programmes (18, 28). At the same time, several of the most frequently detected high-risk genotypes in our study, including HPV31, HPV45, HPV52 and HPV58, are covered by the nonavalent vaccine. These data support ongoing investment in HPV vaccination and suggest that broader genotype coverage of vaccines could provide additional benefit in this population.

HPV infection was associated with condomless sex and the presence of other sexually transmitted infections. Participants reporting condomless sex had more than twice the odds of HPV infection, while those diagnosed with another curable STIs had more than four times the odds of HPV infection. These associations likely reflect shared routes of transmission and common behavioural risk factors (29, 30). The findings support integration of HPV prevention within existing sexual and reproductive health services. While PrEP has become an important component of HIV prevention for AGYW, it does not prevent HPV or other STIs. Condom promotion, STI diagnosis and treatment, contraception services and HPV vaccination therefore remain complementary components of prevention programmes.

The high burden of hrHPV observed in this study presents challenges for cervical cancer screening programmes in rural settings. Clinic-based approaches alone may struggle to achieve adequate coverage, particularly in communities with limited access to healthcare facilities. Self-collected vaginal swabs for HPV testing have shown comparable performance to clinician-collected samples and are generally acceptable to women (31, 32) and were acceptable to women in this rural setting. In high-burden settings such as uMkhanyakude, community-based self-sampling may warrant consideration as a complementary strategy to improve HPV screening coverage in high-burden and underserved communities. Followed by targeted reflexing on cytology or colposcopy among women who tested positive on first-line HPV, with hrHPV, rather than relying solely on centralised facility-based screening.

Further work is needed to understand HPV persistence and immune control in this population. Longitudinal studies are needed to identify factors associated with persistence, clearance and progression to cervical disease, particularly among women living with HIV. Such evidence may also contribute to the development and evaluation of therapeutic HPV vaccines (33).

The strengths of this study include its population-based design, high participation rate and comprehensive HPV genotyping, which enabled assessment of a broad spectrum of HPV types among a representative AGYW sample in an underserved rural community rather than clinic attendees alone. We studied comprehensive HPV genotypes, both low- and high-risk, stratified by HIV status among rural AGYW. Linkage of HPV, HIV and STI data allowed examination of both biological and behavioural correlates of infection. A main limitation of this study is the relatively low response rate for vaginal sample collection. Furthermore, vaginal swab collection differed significantly by HIV status, suggesting the potential for selection bias. As a result, the women who provided vaginal samples may not be fully representative of all women who participated in the survey. Consequently, the findings should be interpreted with caution, as the observed prevalence estimates and associations may not be generalisable to the broader study population. Additionally, the cross-sectional design, which prevents assessment of temporal relationships and persistence, clearance, progression to cervical disease, and the relatively small sample size for subgroup analyses, and the use of age eligibility as a proxy for vaccination status may result in misclassification.

## Conclusion

This population-based study found a high prevalence of hrHPV among AGYW in rural uMkhanyakude. HPV16 and HPV18 were relatively uncommon and were not detected among AGYW eligible for the bivalent HPV vaccine currently used in South Africa’s public programme. In contrast, several of the most common oncogenic genotypes identified are covered by the nonavalent vaccine. These findings support consideration of broader genotype coverage in HPV vaccination programmes, alongside expanded community-based HPV testing as part of cervical cancer screening. Continued surveillance of HPV genotype distribution and longitudinal studies are needed to better understand hrHPV persistence and progression to cervical disease, particularly among women living with HIV.

## Acknowledgents

The authors would like to appreciatively acknowledge the following people:

The *Thethatnami Ngithethenawe* study participants who were all based at uMkhanyakude district, KwaZulu-Natal, South Africa; the Africa Health Research Institute (AHRI) Clinical core study team for recruiting participants and collecting samples in this district; AHRI Biorepository team for their assistance with sample transport to AHRI laboratories, sample distribution and long-term storage of samples and the AHRI Data team for facilitating data collection and data management.

## Authors’ contributions

Conceptualization and methodology, M.M, K.B., Z.N., and M.S.; investigation, M.M, N.Nd. and L.L.; writing - original draft, M.M., and K.B.; writing - review & editing, M.M, K.B., N.Ng. and M.S.; formal analysis, K.B., J.B., and M.M.; funding acquisition, Z.N., and M.S.; resources, N.O., T.K., and C.B.; supervision, K.B., Z.N., and M.S. All authors viewed and approved the final manuscript version.

## Availability of data and materials

The data generated from this study are available upon request to the corresponding author.

## Declarations

### Ethics approval and consent to participate

This study was conducted in accordance with the principles of the Declaration of Helsinki and received ethical approval from the University of KwaZulu-Natal Ethics Committee research ethics policies and regulations. In 2024, ethical approval for the nested study was obtained from the Biomedical Research Ethics Committee at the University of KwaZulu-Natal (BREC/00007429/2024), with expedited approval under the parent protocol (BREC/00003735/2021). Written informed consent was obtained from all participants before study procedures. Participation was voluntary, and participants were informed of their right to withdraw at any time without penalty or impact on their access to healthcare. All procedures were undertaken to minimise potential risks and maximise participant benefits. Participants aged 18 years and above provided written informed consent for the collection of vaginal swabs. For participants younger than 18 years, written parental or guardian consent was obtained, along with assent from the participant.

### Consent for publication

Not applicable.

## Supporting information

Manuscript Tables

## Data Availability

The data generated from this study are available upon request to the corresponding author.

