## Supplementary material for "Prevalence and genotype distribution of human papillomavirus (HPV) among adolescent girls and young women in a high HIV burden rural area of South Africa: a cross-sectional survey": Manuscript Tables

**Table 1. Demographic characteristics of the study sample, overall and by HIV status**

|  | All | AGYW living  with HIV | AGYW Living without HIV | P-value^1^ |
| --- | --- | --- | --- | --- |
| Participants (N) | 187 | 90 | 97 |  |
| Age (Median, IQR) | 26 (22-29) | 26 (22-29) | 25 (21-29) | 0.36^2^ |
| Age groups: (N,%) |  |  |  | 0.64 |
| 15-19 | 16 (8.6 %) | 6 (6.7 %) | 10 (10.3%) |  |
| 20-24 | 61 (32.6%) | 29 (32.2%) | 32 (33.0%) |  |
| 25-30 | 110 (58.8%) | 55 (61.1%) | 55 (56.7%) |  |
| Job status (N,%) |  |  |  | 0.64 |
| Not employed | 136 (72.7%) | 64 (71.1%) | 72 (74.2%) |  |
| Employed | 15 (8.0 %) | 9 (10.0%) | 6 (6.2 %) |  |
| Studying | 36 (19.3%) | 17 (18.9%) | 19 (19.6%) |  |
| Food insecurity in past 12m (N,%) |  |  |  | 0.11 |
| No | 99 (52.9%) | 42 (46.7%) | 57 (58.8%) |  |
| Yes | 88 (47.1%) | 48 (53.3%) | 40 (41.2%) |  |
| Smoked in past 1 month (N,%) |  |  |  | >0.99 |
| No | 181 (96.8%) | 87 (96.7%) | 94 (96.9%) |  |
| Yes | 6 (3.2 %) | 3 (3.3 %) | 3 (3.1 %) |  |
| Alcohol in past 1 month (N,%) |  |  |  | 0.46 |
| No | 151 (81.2%) | 71 (78.9%) | 80 (8283.53%) |  |
| Yes | 35 (18.8%) | 19 (21.1%) | 16 (16.57%) |  |
| Partners in past 12m (N,%) |  |  |  | 0.05 |
| None | 16 (8.6 %) | 4 (4.4 %) | 12 (12.4%) |  |
| 1 | 135 (72.2%) | 72 (80.0%) | 63 (64.9%) |  |
| 2+ | 36 (19.3%) | 14 (15.6%) | 22 (22.7%) |  |
| Sex without condom in past 3m (N,%) |  |  |  | 0.56 |
| No | 89 (48.1%) | 40 (45.5%) | 49 (50.5%) |  |
| Yes | 96 (51.9%) | 48 (54.5%) | 48 (49.5%) |  |
| Using contraception |  |  |  | 0.01 |
| No | 71 (40.8%) | 26 (30.6%) | 45 (50.6%) |  |
| Yes | 103 (59.2%) | 59 (69.4%) | 44 (49.4%) |  |
| Transactional sex in past 12m (N,%) |  |  |  | 0.19 |
| No | 162 (91.5%) | 78 (88.6%) | 84 (94.4%) |  |
| Yes | 15 (8.5 %) | 10 (11.4%) | 5 (5.6 %) |  |
| HIV viral suppression (N,%) |  |  |  | N/A |
| No |  | 40 (44.4%) | N/A |  |
| Yes |  | 50 (55.6%) | N/A |  |
| Any STI infection (N,%) |  |  |  | >0.99 |
| No | 127 (67.9%) | 61 (67.8%) | 66 (68.0%) |  |
| Yes | 60 (32.1%) | 29 (32.2%) | 31 (32.0%) |  |
| Chlamydia (N,%) |  |  |  | 0.61 |
| No | 142 (75.9%) | 70 (77.8%) | 72 (74.2%) |  |
| Yes | 45 (24.1%) | 20 (22.2%) | 25 (25.8%) |  |
| Gonorrhoea (N,%) |  |  |  | 0.38 |
| No | 175 (93.6%) | 86 (95.6%) | 89 (91.8%) |  |
| Yes | 12 (6.4 %) | 4 (4.4 %) | 8 (8.2 %) |  |
| Trichomonas (N,%) |  |  |  | 0.64 |
| No | 167 (89.3%) | 79 (87.8%) | 88 (90.7%) |  |
| Yes | 20 (10.7%) | 11 (12.2%) | 9 (9.3 %) |  |

^1^ Fisher’s exact test comparing HIV negative and HIV positive. ^2^ Wilcoxon rank sum exact p-value comparing HIV negative and HIV positive. The following variables had missing data: alcohol (n=1); sex without condom (n=2); contraceptive use (n=13); transactional sex (n=10).

**Table 2. Prevalence (95% confidence intervals) of HPV DNA by age group and HIV status**

|  | **AGYW living with HIV** | | | | **AGYW without HIV** | | | |
| --- | --- | --- | --- | --- | --- | --- | --- | --- |
|  | **15-19 years (N=6)** | **20-24 years (N=29)** | **25-30 years (N=55)** | **All (N=90)^1^** | **15-19 years (N=10)** | **20-24 years (N=32)** | **25-30 years (N=55)** | **All (N=97)^1^** |
| Any HPV^2^ | 100% | 89.7% (73.6-96.4%) | 78.2% (65.6-87.1%) | 84.6% (75.9-90.6%) | 50.0% (23.7-76.3%) | 81.2% (64.7-91.1%) | 78.2% (65.6-87.1%) | 64.6% (46.6-79.2%) |
| Any HR HPV^3^ | 83.3% (43.6-97.0%) | 62.1% (44.0-77.3%) | 61.8% (48.6-73.5%) | 65.0% (54.3-74.4%) | 50.0% (23.7-76.3%) | 59.4% (42.3-74.5%) | 61.8% (48.6-73.5%) | 55.2% (38.6-70.7%) |
| >1 HR HPV | 16.7% (3.0-56.4%) | 24.1% (12.2-42.1%) | 23.6% (14.4-36.3%) | 22.8% (15.1-32.9%) | 10.0% (1.8-40.4%) | 25.0% (13.3-42.1%) | 34.5% (23.4-47.7%) | 19.6% (10.8-32.9%) |
| Any HR type in 9v vaccine^4^ | 66.7% (30.0-90.3%) | 41.4% (25.5-59.3%) | 41.8% (29.7-55.0%) | 45.3% (34.8-56.2%) | 40.0% (16.8-68.7%) | 31.2% (18.0-48.6%) | 49.1% (36.4-61.9%) | 39.9% (25.3-56.6%) |
| HPV 16/18 | 0 | 24.1% (12.2-42.1%) | 18.2% (10.2-30.3%) | 17.3% (10.9-26.2%) | 0 | 18.8% (8.9-35.3%) | 12.7% (6.3-24.0%) | 7.8 % (4.1 -14.1%) |
| HPV 31/33/45^5^ | 50.0% (18.8-81.2%) | 13.8% (5.5-30.6%) | 12.7% (6.3-24.0%) | 18.4% (10.9-29.4%) | 20.0% (5.7-51.0%) | 6.2% (1.7-20.1%) | 29.1% (18.8-42.1%) | 18.7% (8.9 -35.0%) |

^1^Prevalence weighted to reflect the age distribution of the of the corresponding HIV status target population. ^2^Any of the 38 HPV genotypes detected by the HPV Direct Flow Chip Kit assay: 6, 11, 16, 18, 31, 32, 33, 35, 38, 39, 40, 42, 43, 44, 45, 48, 51, 52, 53, 54, 55, 56, 58, 59, 60, 61, 62, 66, 67, 68, 69, 70, 71, 72, 73, 81, 82 and 84. ^3^HR HPV types in IARC classification Groups 1 or 2a: 16, 18, 31, 33, 35, 39, 45, 51, 52, 56, 58, 59 and 68. ^4^Any of the 7 HR HPV types in the 9-valent vaccine: HPV 16, 18, 31, 33, 45, 52 and 58. ^5^HR HPV types with documented cross-protection by Cervarix

**Table 3. HPV genotype-specific prevalence (95% confidence intervals) by HIV status and overall**

|  | **AGYW living with HIV (N=90)** | | **AGYW without HIV (N=97)** | | **All (N=187)** | |
| --- | --- | --- | --- | --- | --- | --- |
|  | N**^1^** | Prevalence**^2^** | N**^1^** | Prevalence**^2^** | N**^1^** | Prevalence**^3^** |
| High-risk genotypes^4^ |  |  |  |  |  |  |
| HPV16 | 7 | 7.0% (3.4-13.9%) | 6 | 4.4% (1.9-9.9%) | 13 | 5.0% (2.6-9.4%) |
| HPV18 | 10 | 10.2% (5.6-18.0%) | 9 | 4.6% (2.2-9.3%) | 19 | 6.0% (3.5-10.1%) |
| HPV31 | 4 | 5.5% (2.0-14.3%) | 8 | 13.5% (5.0-31.5%) | 12 | 11.6% (4.8-25.5%) |
| HPV33 | 1 | 1.0% (0.2-5.5%) | 6 | 7.2% (2.0-23.2%) | 7 | 5.9% (1.7-18.2%) |
| HPV35 | 17 | 18.8% (11.8-28.6%) | 10 | 5.4% (2.7-10.5%) | 27 | 8.4% (5.2-13.1%) |
| HPV39 | 2 | 2.1% (0.6-7.3%) | 5 | 2.9% (1.1-7.1%) | 7 | 2.8% (1.3-6.3%) |
| HPV45 | 12 | 15.0% (8.4-25.2%) | 11 | 5.5% (2.8-10.4%) | 23 | 7.6% (4.7-12.1%) |
| HPV51 | 6 | 6.1% (2.8-12.8%) | 7 | 4.1% (1.8-8.9%) | 13 | 4.7% (2.5-8.6%) |
| HPV52 | 11 | 11.2% (6.3-19.2%) | 12 | 15.6% (6.5-32.8%) | 23 | 14.6% (7.1-27.6%) |
| HPV56 | 7 | 7.1% (3.5-14.2%) | 9 | 14.3% (5.6-32.0%) | 16 | 12.6% (5.6-26.2%) |
| HPV58 | 11 | 12.6% (6.9-21.8%) | 5 | 2.9% (1.1-7.1%) | 16 | 4.9% (2.7-8.7%) |
| HPV59 | 10 | 11.6% (6.2-20.7%) | 10 | 6.1% (3.0-11.9%) | 20 | 7.4% (4.4-12.3%) |
| HPV68 | 6 | 7.5% (3.3-16.3%) | 9 | 4.6% (2.2-9.3%) | 15 | 5.4% (3.0-9.4%) |
| Low-risk genotypes |  |  |  |  |  |  |
| HPV6 | 8 | 8.1% (4.1-15.3%) | 6 | 3.7% (1.6-8.4%) | 14 | 4.7% (2.5-8.6%) |
| HPV11 | 10 | 10.2% (5.5-17.9%) | 9 | 10.7% (4.1-25.1%) | 19 | 10.6% (4.9-21.2%) |
| HPV32 | 1 | 1.0% (0.2-5.5%) | 0 | ─ | 1 | 0.2% (0.0-1.1%) |
| HPV38 | 1 | 2.4% (0.4-12.5%) | 0 | ─ | 1 | 0.4% (0.1-2.5%) |
| HPV40 | 9 | 9.2% (4.8-16.7%) | 15 | 12.2% (5.2-26.0%) | 24 | 11.8% (5.9-22.0%) |
| HPV42 | 11 | 12.6% (6.9-21.8%) | 7 | 3.7% (1.7-8.2%) | 18 | 5.7% (3.3-9.6%) |
| HPV43 | 5 | 5.0% (2.1-11.4%) | 3 | 1.7% (0.5-5.2%) | 8 | 2.4% (1.1-5.2%) |
| HPV44 | 12 | 12.1% (7.0-20.2%) | 11 | 15.5% (6.5-32.8%) | 23 | 14.6% (7.2-27.6%) |
| HPV48 | 1 | 1.0% (0.2-5.5%) | 0 | ─ | 1 | 0.2% (0.0-1.1%) |
| HPV53 | 2 | 2.0% (0.5-7.2%) | 2 | 0.9% (0.2-3.2%) | 4 | 1.2% (0.4-3.1%) |
| HPV54 | 5 | 5.1% (2.2-11.5%) | 4 | 6.8% (1.7-23.3%) | 9 | 6.3% (2.0-18.3%) |
| HPV55 | 12 | 12.1% (7.0-20.2%) | 11 | 15.5% (6.5-32.8%) | 23 | 14.6% (7.2-27.6%) |
| HPV60 | 0 | ─ | 1 | 0.4% (0.1-2.5%) | 1 | 0.4% (0.1-2.1%) |
| HPV61 | 11 | 11.2% (6.3-19.1%) | 0 | ─ | 11 | 2.3% (1.2-4.3%) |
| HPV62 | 26 | 30.7% (21.5-41.8%) | 16 | 8.7% (4.9-15.1%) | 42 | 13.5% (9.1-19.6%) |
| HPV66 | 3 | 3.1% (1.0-8.7%) | 2 | 0.9% (0.2-3.2%) | 5 | 1.4% (0.5-3.4%) |
| HPV67 | 4 | 5.5% (2.0-14.3%) | 5 | 11.9% (3.9-30.7%) | 9 | 10.2% (3.8-24.7%) |
| HPV69 | 1 | 1.0% (0.2-5.7%) | 2 | 1.2% (0.3-4.7%) | 3 | 1.2% (0.4-4.0%) |
| HPV70 | 6 | 7.5% (3.3-16.4%) | 5 | 2.9% (1.1-7.1%) | 11 | 3.9% (2.0-7.5%) |
| HPV71 | 15 | 16.7% (10.1-26.3%) | 8 | 4.2% (1.9-8.7%) | 23 | 6.9% (4.2-11.1%) |
| HPV72 | 4 | 5.5% (2.0-14.3%) | 1 | 0.8% (0.1-4.5%) | 5 | 1.7% (0.7-4.5%) |
| HPV73 | 4 | 5.4% (2.0-14.2%) | 1 | 0.8% (0.1-4.5%) | 5 | 1.7% (0.6-4.5%) |
| HPV81 | 26 | 30.7% (21.5-41.8%) | 16 | 8.7% (4.9-15.1%) | 42 | 13.5% (9.1-19.6%) |
| HPV82 | 3 | 3.1% (1.0-8.8%) | 3 | 6.0% (1.3-23.4%) | 6 | 5.3% (1.4-18.2%) |
| HPV84 | 3 | 3.1% (1.0-8.7%) | 2 | 1.2% (0.3-4.7%) | 5 | 1.6% (0.6-4.3%) |

^1^Number of participants with genotype-specific DNA detected. ^2^Prevalence weighted to reflect the age distribution of the population. ^3^Prevalence weighted to reflect the joint age and HIV status distribution of the population. ^4^HR HPV types in IARC classification Groups 1 or 2a: 16, 18, 31, 33, 35, 39, 45, 51, 52, 56, 58, 59 and 68.

**Table 4. Factors associated with any HPV DNA**

|  | **n with any HPV DNA/ N (%)^1^** | **Adjusted OR (95% CI)^2^** |
| --- | --- | --- |
| Age group (years) |  | P=0.30^3^ |
| 15-19 | 11 / 16 (68.8%) | 1 |
| 20-24 | 55 / 64 (85.9%) | 2.54 (0.74 -8.70) |
| 25-30 | 88 / 112 (78.6%) | 1.60 (0.53 -4.89) |
| HIV status |  | P=0.25 |
| Not living with HIV | 74 / 97 (76.3%) | 1 |
| Living with HIV | 75 / 90 (83.3%) | 1.51 (0.74 -3.11) |
| Job status |  | P=0.12 |
| Not employed | 107 / 136 (78.7%) | 1 |
| Employed | 10 / 15 (66.7%) | 0.50 (0.16 -1.58) |
| Studying | 32 / 36 (88.9%) | 2.35 (0.76 -7.32) |
| Food insecurity in past 12m |  | P=0.77 |
| No | 78 / 99 (78.8%) | 1 |
| Yes | 71 / 88 (80.7%) | 1.11 (0.54 -2.29) |
| Alcohol in past 1 month |  | P=0.04 |
| No | 124 / 151 (82.1%) | 1 |
| Yes | 24 / 35 (68.6%) | 0.40 (0.17 -0.94) |
| Partners in past 12m |  | P=0.07 |
| None | 9 / 16 (56.2%) | 1 |
| 1 | 108 / 135 (80.0%) | 2.58 (0.83 -7.98) |
| 2+ | 32 / 36 (88.9%) | 5.19 (1.25 -21.58) |
| Sex without condom in past 3m |  | P=0.03 |
| No | 65 / 89 (73.0%) | 1 |
| Yes | 83 / 96 (86.5%) | 2.23 (1.06 -4.70) |
| Transactional sex in past 12m |  | P=0.25 |
| No | 130 / 162 (80.2%) | 1 |
| Yes | 14 / 15 (93.3%) | 2.50 (0.44 -14.12) |
| Any STI infection |  | P=0.002 |
| No | 93 / 127 (73.2%) | 1 |
| Yes | 56 / 60 (93.3%) | 4.31 (1.52 -12.20) |
| Chlamydia |  | P=0.04 |
| No | 108 / 142 (76.1%) | 1 |
| Yes | 41 / 45 (91.1%) | 2.80 (0.98 -8.00) |
| Gonorrhoea |  | P=0.06 |
| No | 137 / 175 (78.3%) | 1 |
| Yes | 12 / 12 (100.0%) | 7.68 (0.44 -134.02) |
| Trichomonas |  | P=0.11 |
| No | 130 / 167 (77.8%) | 1 |
| Yes | 19 / 20 (95.0%) | 3.34 (0.61 -18.44) |

^1^Unweighted proportions. ^2^Adjusted for age group and HIV status. ^3^P-values from penalised likelihood ratio test.
