## Supplementary Tables for "Prevalence and genotype distribution of human papillomavirus (HPV) among adolescent girls and young women in a high HIV burden rural area of South Africa: a cross-sectional survey"

**Supplementary Table 1:** **Participant flow for STI testing consent and specimen collection by HIV status**

|  | **WLWH (n=214)** | **WLWoH (n=811)** | **Total (n=1,025)** |
| --- | --- | --- | --- |
| **Consenting to STI testing** | | | |
| Consented to STI testing | 171 (79.9%) | 608 (75.0%) | 779 (76.0%) |
| *Did not consent to STI testing* | *43 (20.1%)* | *203 (25.0%)* | *246 (24.0%)* |
| **Specimen collection** | | | |
| **No STI specimen collected** | 7/171 (4.1%) | 36/608 (5.9%) | 43/779 (5.5%) |
| **STI specimen collected** | 164/171 (95.9%) | 572/608 (94.1%) | 736/779 (94.5%) |
| **Type of specimen collected** | | | |
| **Vaginal swab collected** | **101/164 (61.6%)** | **282/572 (49.3%)** | **383/736 (52.0%)** |
| **Urine sample collected** | 63/164 (38.4%) | 290/572 (50.7%) | 353/736 (48.0%) |

HIV status was determined from dried blood spot (DBS) samples collected during the same survey visit. STI specimens were self-collected during the survey, and participants could choose to provide either a vaginal swab or a urine sample.

**Supplementary Table 2:** **Association between STI specimen type and HIV status**

| **Characteristic** | **Urine, n/N (%)** | **Vaginal swab, n/N (%)** | **Crude OR (95% CI)** | **Adjusted OR (95% CI)** | **p-value** |
| --- | --- | --- | --- | --- | --- |
| WLWoH | 290/572 (50.7%) | 282/572 (49.3%) | Ref | Ref |  |
| WLWH | 63/164 (38.4%) | 101/164 (61.6%) | 1.70 (1.20, 2.41) | 1.30 (0.89, 1.90) | 0.177 |
| No STI symptom | 189/350 (54.0%) | 161/350 (46.0%) | Ref | Ref |  |
| Any STI symptom | 164/386 (42.5%) | 222/386 (57.5%) | 1.60 (1.20, 2.13) | 1.44 (1.07, 1.95) | 0.016 |
| 15-19 | 145/239 (60.7%) | 94/239 (39.3%) | Ref | Ref |  |
| 20-24 | 104/246 (42.3%) | 142/246 (57.7%) | 2.12 (1.49, 3.02) | 1.42 (0.91, 2.21) | 0.123 |
| 25-30 | 104/251 (41.4%) | 147/251 (58.6%) | 2.09 (1.47, 2.97) | 1.24 (0.75, 2.06) | 0.402 |
| Not in school | 205/489 (41.9%) | 284/489 (58.1%) | Ref | Ref |  |
| Currently in school | 147/246 (59.8%) | 99/246 (40.2%) | 0.47 (0.35, 0.63) | 0.61 (0.40, 0.94) | 0.026 |
| Currently pregnant | 15/39 (38.5%) | 24/39 (61.5%) | 1.62 (0.84, 3.10) | 1.44 (0.74, 2.80) |  |
| Not pregnant | 328/667 (49.2%) | 339/667 (50.8%) | Ref | Ref | 0.279 |

Note: Outcome is vaginal swab versus urine sample. Percentages are calculated within each category. Crude and adjusted ORs are from GEE logistic regression models. Adjusted model includes HIV status, STI symptoms, age group, school status and current pregnancy. ORs greater than 1 indicate higher odds of providing a vaginal swab.

**Supplementary Table 3: Prevalence (95% confidence intervals) of HPV DNA by age group^1^**

|  | **15-19 years (n=16)** | **20-24 years (n=61)** | **25-30 years (n=110)** | **All (n=187)** |
| --- | --- | --- | --- | --- |
| Any HPV^2^ | 53.2% (27.0-77.7%) | 83.0% (69.5-91.3%) | 78.2% (69.2-85.1%) | 69.1% (54.5-80.7%) |
| Any HR HPV^3^ | 52.1% (26.2-76.9%) | 60.0% (45.7-72.7%) | 61.8% (52.1-70.7%) | 57.3% (44.1-69.6%) |
| >1 HR HPV | 10.4% (2.1 -38.6%) | 24.8% (14.7-38.8%) | 30.5% (22.3-40.2%) | 20.7% (13.2-30.9%) |
| Any HR type in 9v vaccine^4^ | 41.7% (18.8-68.8%) | 33.4% (21.7-47.5%) | 46.4% (37.0-56.1%) | 41.0% (29.2-54.1%) |
| HPV 16/18 | 0 | 19.9% (11.1-33.2%) | 14.7% (9.2 -22.8%) | 10.0% (6.4 -15.4%) |
| HPV 31/33/45^5^ | 21.9% (7.2 -50.5%) | 7.9 % (3.2 -18.2%) | 23.1% (15.8-32.5%) | 18.6% (10.4-30.9%) |

^1^Prevalence weighted to reflect the joint age and HIV status distribution of the population. ^2^Any of the 38 HPV genotypes detected by the HPV Direct Flow Chip Kit assay: 6, 11, 16, 18, 31, 32, 33, 35, 38, 39, 40, 42, 43, 44, 45, 48, 51, 52, 53, 54, 55, 56, 58, 59, 60, 61, 62, 66, 67, 68, 69, 70, 71, 72, 73, 81, 82 and 84. ^3^HR HPV types in IARC classification Groups 1 or 2a: 16, 18, 31, 33, 35, 39, 45, 51, 52, 56, 58, 59 and 68. ^4^Any of the 7 HR HPV types in the 9-valent vaccine: HPV 16, 18, 31, 33, 45, 52 and 58. ^5^HR HPV types with documented cross-protection by Cervarix

**Supplementary table 4: Prevalence (95% confidence intervals) of HPV DNA by whether likely to have been offered HPV vaccination**

|  | **Age 15-19 years** | | | **Age 20-30 years** | | |
| --- | --- | --- | --- | --- | --- | --- |
|  | **AGYW living with HIV (N=6)** | **AGYW without HIV (N=10)** | **All  (N=16)^1^** | **AGYW living with HIV (N=84)^2^** | **AGYW without HIV (N=87)^2^** | **All  (N=171)^3^** |
| Any HPV^4^ | 100.0% | 50.0% (23.3-76.7%) | 53.2% (27.0-77.7%) | 82.0% (72.2-88.9%) | 79.8% (69.5-87.2%) | 80.4% (72.9-86.2%) |
| Any HR HPV^5^ | 83.3% (43.0-97.1%) | 50.0% (23.3-76.7%) | 52.1% (26.2-76.9%) | 61.9% (51.0-71.7%) | 60.6% (49.3-70.8%) | 61.0% (52.6-68.8%) |
| >1 HR HPV | 16.7% (2.9 -57.0%) | 10.0% (1.7 -41.0%) | 10.4% (2.1 -38.6%) | 23.8% (15.8-34.1%) | 29.6% (20.7-40.4%) | 28.0% (21.1-36.0%) |
| Any HR type in 9v vaccine^6^ | 66.7% (29.5-90.5%) | 40.0% (16.6-69.1%) | 41.7% (18.8-68.8%) | 41.7% (31.5-52.6%) | 39.8% (29.7-50.9%) | 40.6% (32.8-48.9%) |
| HPV 16/18 | 0 | 0 | 0 | 20.2% (12.9-30.2%) | 15.8% (9.2 -25.9%) | 17.1% (11.7-24.2%) |
| HPV 31/33/45^7^ | 50.0% (18.4-81.6%) | 20.0% (5.6 -51.5%) | 21.9% (7.2 -50.5%) | 13.1% (7.4 -22.1%) | 17.3% (10.8-26.3%) | 16.2% (11.2-22.9%) |

^1^Prevalence weighted to reflect the HIV distribution of the target population aged 15-19 years. ^2^Prevalence weighted to reflect the age distribution of the corresponding HIV status target population. ^3^Prevalence weighted to reflect the joint age and HIV distribution of the target population. ^4^Any of the 38 HPV genotypes detected by the HPV Direct Flow Chip Kit assay: 6, 11, 16, 18, 31, 32, 33, 35, 38, 39, 40, 42, 43, 44, 45, 48, 51, 52, 53, 54, 55, 56, 58, 59, 60, 61, 62, 66, 67, 68, 69, 70, 71, 72, 73, 81, 82 and 84. ^5^HR HPV types in IARC classification Groups 1 or 2a: 16, 18, 31, 33, 35, 39, 45, 51, 52, 56, 58, 59 and 68. ^6^Any of the 7 HR HPV types in the 9-valent vaccine: HPV 16, 18, 31, 33, 45, 52 and 58. ^7^HR HPV types with documented cross-protection by Cervarix.
